# Echocardiographic Characteristics and Non-Dipping Blood Pressure Profile: Is There an Association?

**DOI:** 10.1101/2024.05.23.24307846

**Authors:** Daniel Villarreal, Federico Ramos, Daniel Gasca, Camilo Roa, Mariangela Cardone, Carolina Ayala, Camilo Alvarado

## Abstract

**Background:** The circadian rhythm of blood pressure is a fundamental aspect of cardiovascular physiology in healthy individuals. Beyond nocturnal hypertension, a blunted or impaired BP circadian variation is linked to heightened target organ damage and elevated cardiovascular disease risk. This includes alterations in cardiac structure and function, atherosclerotic cardiovascular disease, dementia and heart failure.

**Methods:** A retrospective cohort study involving 1600 participants enrolled between 2021 and 2023 identified 847 as dippers and 753 as non-dippers based on 24-hour ambulatory blood pressure monitoring. Echocardiographic evaluations were performed to assess cardiac structure and function.

**Results:** Non-dipping individuals displayed more signs of adverse cardiac remodeling, including a higher rate of eccentric hypertrophy (1.73 vs. 0.47%), increased left ventricular mass index in both men (75.82 vs. 70.10 g/m^2^) and women (65.44 vs. 63.92 g/m^2^), left ventricular internal diameter in diastole (4.38 vs. 4.23 cm), and left ventricular posterior wall thickness (0.82 vs. 0.81 cm). Additionally, non-dipping participants exhibited impaired ventricular relaxation, with higher E/e’ ratios medially (9.45 vs. 8.86) and laterally (7.61 vs. 7.23) and rates of type 1 diastolic dysfunction (9.31 vs. 4.49%). These differences persisted when analysing only participants with hypertension.

**Conclusions:** Our study highlights the substantial impact of non-dipping blood pressure patterns on cardiac structure and function. It suggests that non-dipping blood pressure patterns may serve as an independent predictor of adverse cardiac remodelling, irrespective of hypertension diagnosis. These results underscore the necessity of devising monitoring strategies and implementing targeted interventions to address the cardiovascular risks associated with non-dipping BP profile.

**Graphical abstract:** 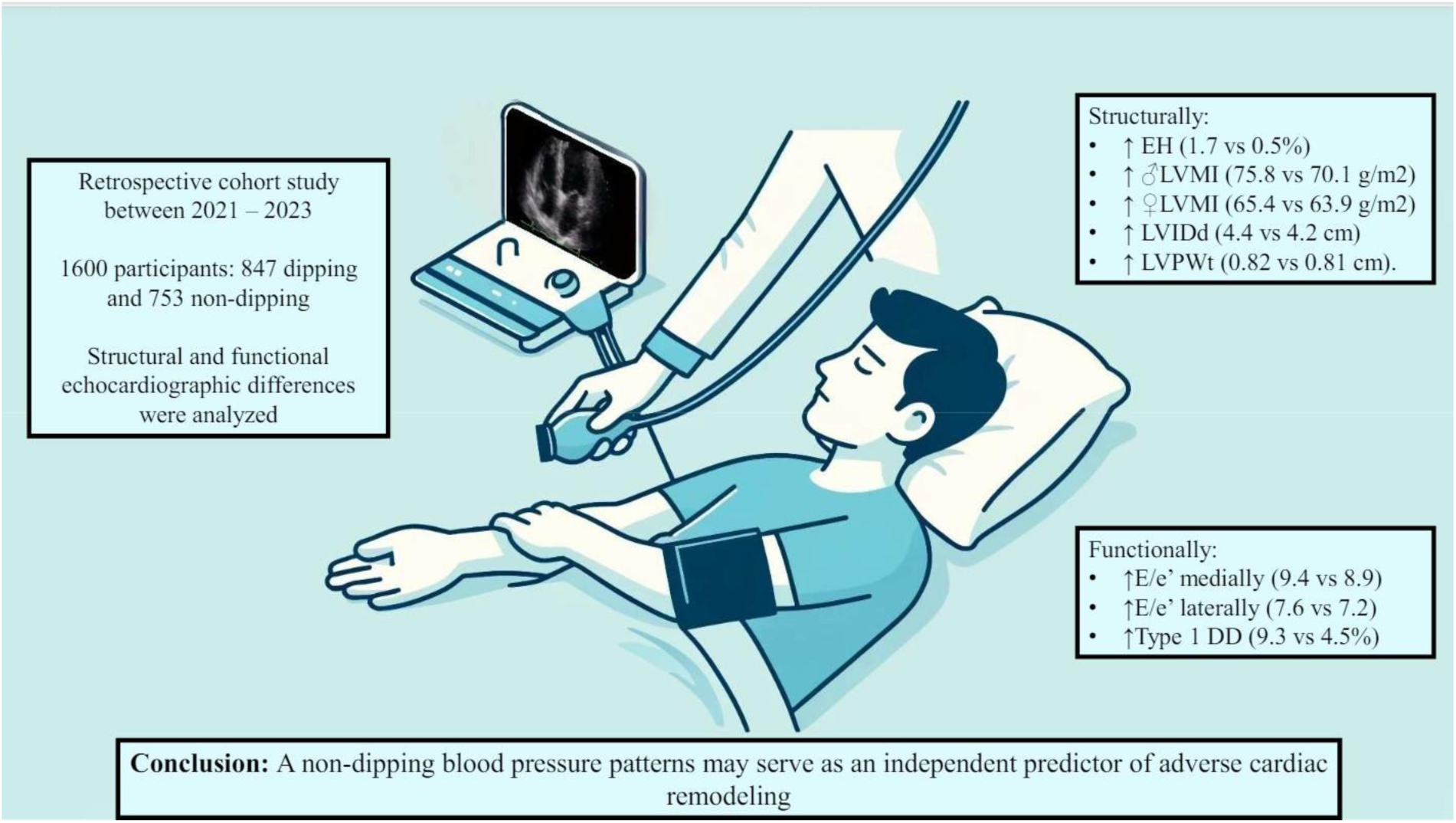

## Introduction

The circadian rhythm of blood pressure (BP) is a fundamental aspect of cardiovascular physiology in healthy individuals. It is characterized by fluctuations throughout the day and typically, there is a nocturnal dip of 10 – 20% from mean daytime BP levels, accompanied by a pronounced morning surge upon awakening.^1^

24-hour Ambulatory Blood Pressure Monitoring (ABPM) has emerged as a valuable tool for recording and monitoring a patient’s nighttime to daytime BP ratio, enabling classification based on their nocturnal dip pattern. Patients can be categorized as dippers, exhibiting a normal physiologic circadian rhythm; non-dippers, with a nighttime BP dip ranging from 0 to 10% of their daytime average; extreme dippers, characterized by a nighttime BP dip exceeding 20%; and reverse dippers, wherein nocturnal BP surpasses daytime levels.^2^

An impaired circadian BP response has been consistently linked to elevated risks of mortality, dementia, and adverse cardiovascular and renal outcomes.^3–8^ Even in the absence of nocturnal hypertension, a blunted or impaired BP circadian variation has been associated with increased target organ damage and risk of cardiovascular disease, including alterations in cardiac structure and function, atherosclerotic cardiovascular disease, and heart failure.^9^

The sustained and persistent elevation in nocturnal blood pressure exerts cumulative effects on cardiac structure and function over the long term, including left ventricular hypertrophy, impaired diastolic and systolic function, changes in left ventricular geometry, and enlargement of the left atrium, among others.^10,11^ Research indicates that a non-dipping pattern observed in individuals with primary arterial hypertension correlates with increased relative wall thickness (RWT), left ventricular mass index (LVMI), and left atrial diameter (LAD), as well as reduced E/A ratio, isovolumetric relaxation time (IRT), myocardial performance index (MPI), and peak early and late myocardial diastolic velocity.^12,13^

However, existing studies predominantly focus on patients with uncontrolled hypertension. There remains a dearth of evidence regarding the impact of a non-dipping pattern on cardiac function and structure, irrespective of hypertension diagnosis. Furthermore, scant evidence exists within the Colombian and Latin American populations regarding echocardiographic findings associated with this circadian alteration in blood pressure. Consequently, we conducted a retrospective cohort study to investigate the echocardiographic changes linked with a non-dipping pattern of blood pressure in both normotensive and hypertensive individuals.

## Methods

We conducted a retrospective cohort study involving 1600 participants enrolled between 2021 and 2023. A comprehensive outline of the methodology is available in <u>Supplemental Methods</u>, while a condensed overview is presented herein. Please see the Major Resources Table in the Supplemental Materials. The study adhered to the ethical principles delineated in the Declaration of Helsinki, as well as local and international standards governing ethical research. Approval was obtained from the Institutional Review Board (IRB) of Fundación Santa Fe de Bogotá prior to commencement of data collection. As per local regulations, informed consent was not required due to the absence of interventions, anonymization of data, and sole reliance on medical record review.

### Study population

Participants older than 18 years of age who had a dipping or non-dipping pattern on 24-hour ABPM performed between the years 2021 and 2023 and an echocardiogram within 2 years of the ABPM, were included. Exclusion criteria encompassed patients who had their 24-hour ABPM or echocardiogram performed as an inpatient or in the context of an acute or acute-on-chronic condition. A full list of the inclusion and exclusion criteria is available in <u>Supplementary File S1</u>.

Participants were selected from a database of 11,425 patients who had a 24-hour ABPM performed between the years 2021 and 2023 at our institution. After eliminating duplicates and filtering for those who had an echocardiogram performed within 2 years of the 24-hour ABMP, 5,404 participants remained. Of these, 2,776 had a dipping pattern, 1,275 were non-dipping, 1,104 had extreme dipping and 249 had an inverse dipping pattern. Using simple randomization, 1,600 participants were picked for final analysis.

### Data collection

Sociodemographic, clinical, echocardiographic and ABPM data were collected from the participants’ clinical records by the investigators and stored in a confidential database accessible only to them. The 24-hour ABPM device (*Labsa IonTrack 90217a-1*) was fitted, and patients were instructed about its use by a trained-nurse and readings were interpreted by a cardiologist. 2-dimensional transthoracic echocardiography (*General Electric* or *Philips*) was performed by a certified sonographer using the recommendations of the American Society of Echocardiography (ASE) and results interpreted by a cardiologist specialized in diagnostic imaging.^14^ Neither of the cardiologist who interpreted the 24-hour ABPM nor the echocardiography were involved in the study.

Clinical, anthropometric and sociodemographic characteristics were obtained from medical records of outpatient visits. The primary outcome was structural and functional differences in echocardiographic parameters between dippers and non-dippers.

### Statistical Analysis

A comprehensive descriptive analysis encompassing participants sociodemographic, anthropometric, clinical, echocardiographic, and ABPM characteristics was conducted. Subsequently, in accordance with established literature and considering the robustness of the sample size and the fulfilment of assumptions prescribed by the Central Limit Theorem, the need for normality testing was deemed unnecessary.^15–17^

Echocardiographic parameters were assessed for disparities between individuals categorized as dippers and non-dippers using independent Student’s t-test, Analysis of Covariance (ANCOVA), Pearson’s χ2 test, or Chi-squared trend test, contingent upon the inherent characteristics and distribution of the data. Adjustments for age, sex, diagnosis of heart failure, duration of usage of antihypertensive medications, and mean arterial 24-hour blood pressure were implemented. Subsequently, subgroup analyses were performed employing the same analytical framework, but stratifying based on the diagnosis of hypertension prior to the 24-hour ABPM. A two-tailed alpha error of 5% and a statistical power of 80% were used to ascertain statistical significance. DV as principal author had full access to all the data in the study and takes responsibility for its integrity and the data analysis.

## Results

### Baseline characteristics

Among the patients included in our study, 847 exhibited a dipping pattern on 24-hour ABPM, while 753 had a non-dipping pattern, roughly representing a ratio of 1.1 to 1. Overall, 53.4% of the participants were female, with a mean age of 64.4 years and a standard deviation of 14.46. Both groups showed a similar distribution of sex, height, weight, and body mass index (BMI) compared to the overall population. However, non-dipping participants were older than those in the dipping group (*p* <0.01), consistent with findings from several studies indicating that aging is associated with a non-dipping BP profile, likely due to changes in the aging cardiovascular system.^18^

Interestingly, participants in the non-dipping group also exhibited an increased prevalence of hypertension (58.8% vs. 52.3%), heart failure (50.5% vs. 28.3%), and coronary artery disease (13.8% vs. 9.7%) compared to individuals with a dipping BP. This, along with the overall higher mean systolic and diastolic BP readings on 24-hour ABPM, may contribute to the higher likelihood of patients in the non-dipping group being prescribed an antihypertensive medication (59.4% vs. 49.2%). Nonetheless, the duration of antihypertensive medication use did not differ statistically between the two groups (Table 1).

**Table 1.**
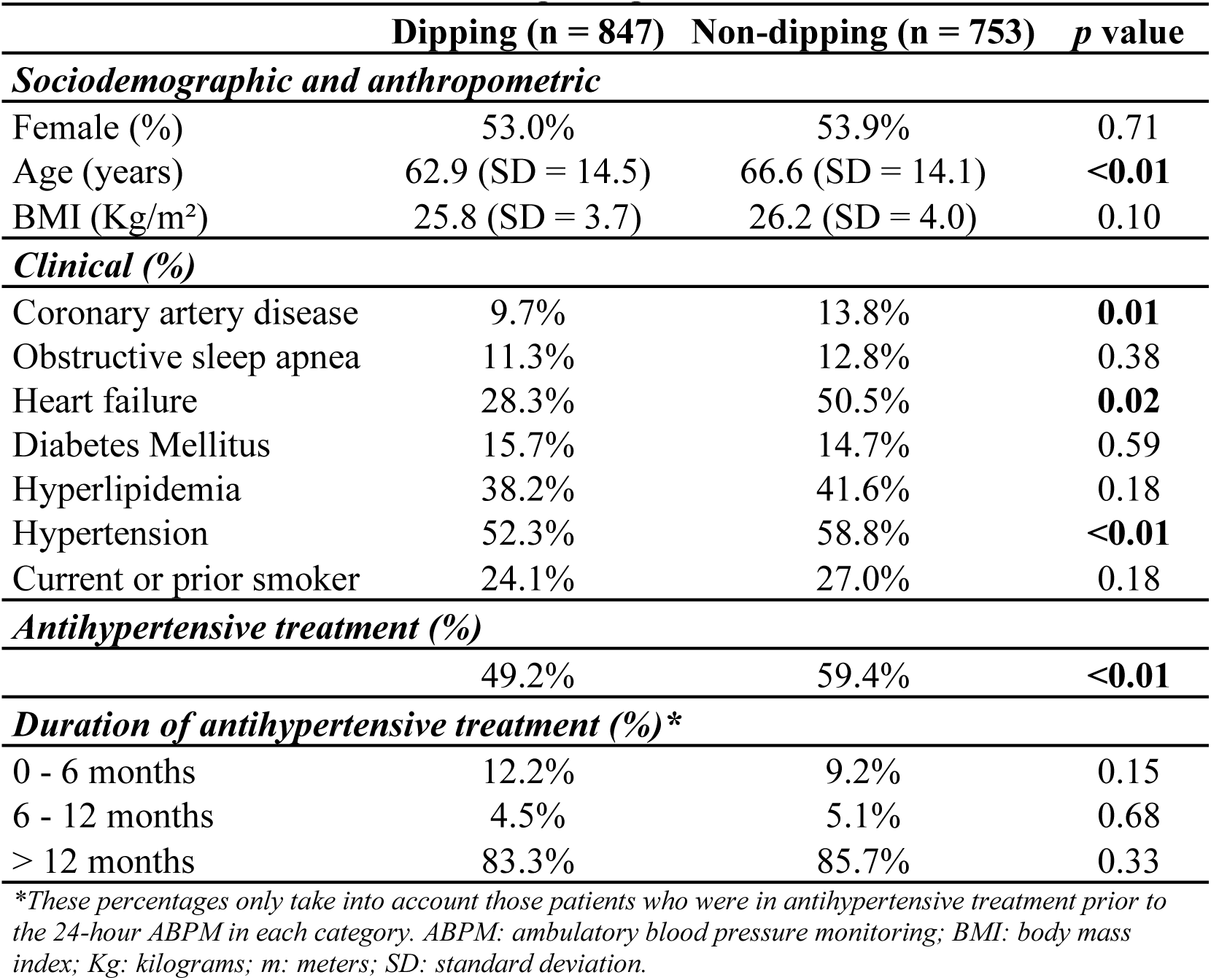
Baseline characteristics of participants.

|  | **Dipping (n = 847)** | **Non-dipping (n = 753)** | **p value** |
| --- | --- | --- | --- |
| ***Sociodemographic and anthropometric*** |  |  |  |
| Female (%) | 53.0% | 53.9% | 0.71 |
| Age (years) | 62.9 (SD = 14.5) | 66.6 (SD = 14.1) | **<0.01** |
| BMI (Kg/m<sup>2</sup>) | 25.8 (SD = 3.7) | 26.2 (SD = 4.0) | 0.10 |
| ***Clinical (%)*** |  |  |  |
| Coronary artery disease | 9.7% | 13.8% | **0.01** |
| Obstructive sleep apnea | 11.3% | 12.8% | 0.38 |
| Heart failure | 28.3% | 50.5% | **0.02** |
| Diabetes Mellitus | 15.7% | 14.7% | 0.59 |
| Hyperlipidemia | 38.2% | 41.6% | 0.18 |
| Hypertension | 52.3% | 58.8% | **<0.01** |
| Current or prior smoker | 24.1% | 27.0% | 0.18 |
| ***Antihypertensive treatment (%)*** |  |  |  |
|  | 49.2% | 59.4% | **<0.01** |
| ***Duration of antihypertensive treatment (%)\**** |  |  |  |
| 0 - 6 months | 12.2% | 9.2% | 0.15 |
| 6 - 12 months | 4.5% | 5.1% | 0.68 |
| > 12 months | 83.3% | 85.7% | 0.33 |

\*These percentages only take into account those patients who were in antihypertensive treatment prior to the 24-hour ABPM in each category. ABPM: ambulatory blood pressure monitoring; BMI: body mass index; Kg: kilograms; m: meters; SD: standard deviation.

### 24-hour ABPM

As expected, patients in the non-dipping group exhibited significantly higher mean systolic and diastolic pressures during nighttime, likely contributing to the increased percentage of systolic blood pressure readings above the upper normal limit, as indicated by the systolic BP load. Additionally, they demonstrated significantly higher mean 24-hour systolic and daytime diastolic blood pressure readings, suggesting a potentially more severe degree of hypertension in this group (Table 2). Furthermore, non-dipping participants displayed a higher ambulatory arterial stiffness index (AASI). This finding is consistent with previous studies that have shown an inverse association between AASI and systolic dipping, and a direct one with a non-dipping profile.^19^ The observed association between AASI and a non-dipping profile may also be influenced by the older age of the non-dipping participants in our study.

**Table 2.** 24-hour ABPM characteristics.

|  | **Dipping**<br>**(n = 847)** | **Non-dipping**<br>**(n = 753)** | ***p* value** |
| --- | --- | --- | --- |
| 24-hour MSP (mmHg) | 115.8 | 118.3 | **<0.01** |
| 24-hour MDP (mmHg) | 70.8 | 80.0 | 0.74 |
| Daytime MSP (mmHg) | 121.2 | 121.2 | 0.95 |
| Daytime MDP (mmHg) | 75.0 | 73.3 | **<0.01** |
| Nighttime MSP (mmHg) | 104.7 | 112.3 | **<0.01** |
| Nighttime MDP (mmHg) | 62.5 | 66.3 | **<0.01** |
| Systolic BP load (%) | 18.7 | 23.7 | **<0.01** |
| Diastolic BP load (%) | 20.9 | 22.4 | 0.09 |
| AASI | 0.42 | 0.50 | **0.01** |
| Mean systolic nighttime dipping (%) | 13.6 | 7.3 | **<0.01** |
| Mean diastolic nighttime dipping (%) | 16.6 | 9.6 | **<0.01** |

*AASI: ambulatory arterial stiffness index; ABPM: ambulatory blood pressure monitor; BP: blood pressure; MDP: mean diastolic pressure; MSP: mean systolic pressure.*

### Non-dipping BP profile and echocardiographic findings in the overall population

In the overall population, both men and women with a non-dipping BP profile displayed an increased LVMI alongside a normal RWT, suggestive of eccentric cardiac hypertrophy. This observation highlights the potential role of non-dipping BP profile in the development of this type of remodelling, given its heightened prevalence in this group. Factors such as increased circulating plasma volume and preload, characteristic of the non-dipping BP profile, likely contribute to this association.^20^ Furthermore, the finding of an increased left ventricular internal diameter in diastole (LVIDd) and left ventricular posterior wall (LVPW) thickness in the non-dipping group provides additional support for adverse cardiac remodelling in this population.

Functionally, non-dipping participants exhibited a decreased left ventricular ejection fraction (LVEF) compared to their dipping counterparts. Tissue Doppler analysis revealed higher E/e’ ratios, both medially and laterally, in the non-dipping group. While the E/A ratio was similar in both groups, the elevated E/e’ ratio suggests impaired ventricular relaxation, which may progress to overt diastolic dysfunction. This finding is further supported by the increased prevalence of diastolic dysfunction observed in the non-dipping group. Participants from both groups tended to have milder forms of diastolic dysfunction, although only the non-dipping group included participants with reversible and non-reversible restrictive diastolic dysfunction.

Right sided heart functional and structural assessment didn’t show any significant differences between both groups (Table 3 and Supplemental Table S1).

**Table 3.** Echocardiographic parameters in the overall patient population*.

|  | Dipping (n = 847) | Non-dipping (n = 753) | p value |
| --- | --- | --- | --- |
| ***Structural assessment*** |  |  |  |
| LVMI (g/m<sup>2</sup>) |  |  |  |
| Men | 70.1 | 75.82 | **<0.01** |
| Women | 63.92 | 65.44 | 0.18 |
| RWT | 0.39 | 0.38 | 0.07 |
| IVS thickness (cm) | 0.89 | 0.91 | 0.06 |
| LVPW thickness (cm) | 0.81 | 0.82 | **0.03** |
| LVIDs (cm) | 2.81 | 2.82 | 0.92 |
| LVIDd (cm) | 4.23 | 4.38 | **<0.01** |
| LAVI (mL/m<sup>2</sup>) | 25.85 | 26.33 | 0.27 |
| Cardiac hypertrophy (%) |  |  |  |
| Concentric | 14.64 | 13.81 | 0.64 |
| Excentric | 0.47 | 1.73 | **0.01** |
| ***Functional assessment*** |  |  |  |
| LVEF (%) | 66.2 | 61.6 | **<0.01** |
| E/A | 0.92 | 0.93 | 0.81 |
| E/e' septal | 8.86 | 9.45 | **<0.01** |
| E/e' lateral | 7.23 | 7.61 | **0.02** |
| Dyastolic dysfunction (%) |  |  |  |
| Grade 1 | 4.49 | 9.31 | **<0.01** |
| Grade 2 | 0.93 | 1.06 | 0.63 |
| Grade 3 | 0 | 1.06 | - |
| Grade 4 | 0 | 0.13 | - |
| ***Right chambers and pulmonary circulation assessment*** |  |  |  |
| RVIDd (cm) | 3.32 | 3.36 | 0.09 |
| TAPSE (mm) | 22.30 | 22.40 | 0.58 |
| PASP (mmHg) | 31.83 | 31.92 | 0.86 |

*These analyses included hypertensive and non-hypertensive patients and were adjusted by age, sex, BMI, heart failure, use and duration of antihypertensive treatment and 24-hour MAP. IVS: interventricular septum; LAVI: left atrial volume index; LVIDd: left ventricular internal diameter in diastole; LVIDs: left ventricular internal diameter in systole; LVMI: left ventricular mass index; LVPW: left ventricle posterior wall; MAP: mean arterial pressure; PASP: pulmonary artery systolic pressure; RVIDd: right ventricular internal diameter in diastole; RWT: relative wall thickness; TAPSE: tricuspid annular plane systolic excursion.*

### Non-dipping BP profile and echocardiographic findings in the hypertensive population

Hypertensive participants with a non-dipping profile exhibited increased LVMI, LVIDd and lower RWT compared to those with hypertension and a normal dipping profile. Although the proportion of patients with eccentric hypertrophy was numerically higher in the non-dipping group, there were no statistically significant differences between the two groups. While the lack of statistical significance may be attributed to the study’s power limitations, these findings suggest that a non-dipping profile could serve as an independent risk factor for adverse cardiac remodelling, even in hypertensive patients.

Functional assessment revealed an increased percentage of patients with type 1 diastolic dysfunction and reversible restrictive diastolic dysfunction in the non-dipping group. Additionally, other variables related to impaired ventricular relaxation showed higher values in the non-dipping group, although they did not reach statistical significance. Conversely, right-sided heart functional and structural assessment did not show any significant differences between both groups (Table 4 and Supplemental Table S2).

**Table 4.** Echocardiographic parameters in the hypertensive patient population*.

|  | Dipping (n = 442) | Non-dipping (n = 443) | p value |
| --- | --- | --- | --- |
| ***Structural assessment*** |  |  |  |
| LVMI (g/m<sup>2</sup>) |  |  |  |
| Men | 72.32 | 77.34 | **0.02** |
| Women | 66.00 | 69.34 | **0.03** |
| RWT | 0.40 | 0.39 | **0.02** |
| IVS thickness (cm) | 0.93 | 0.94 | 0.19 |
| LVPW thickness (cm) | 0.82 | 0.84 | 0.18 |
| LVIDs (cm) | 2.81 | 2.83 | 0.82 |
| LVIDd (cm) | 4.19 | 4.36 | **<0.01** |
| LAVI (mL/m<sup>2</sup>) | 27.30 | 27.74 | 0.48 |
| Cardiac hypertrophy (%) |  |  |  |
| Concentric | 20.59 | 18.06 | 0.34 |
| Excentric | 0.90 | 2.03 | 0.16 |
| ***Functional assessment*** |  |  |  |
| LVEF (%) | 63.00 | 62.24 | 0.06 |
| E/A | 0.88 | 0.88 | 0.98 |
| E/e' septal | 9.50 | 9.99 | 0.06 |
| E/e' lateral | 7.84 | 8.03 | 0.38 |
| Dyastolic dysfunction (%) |  |  |  |
| Grade 1 | 6.11 | 12.44 | **<0.01** |
| Grade 2 | 1.58 | 1.36 | 0.78 |
| Grade 3 | 0 | 1.13 | - |
| Grade 4 | 0 | 0 | - |
| ***Right chambers and pulmonary circulation assessment*** |  |  |  |
| RVIDd (cm) | 3.31 | 3.36 | 0.10 |
| TAPSE (mm) | 22.02 | 22.27 | 0.30 |
| PASP (mmHg) | 32.91 | 33.16 | 0.73 |

*These analyses only included hypertensive patients and were adjusted by age, sex, BMI, heart failure, type and duration of antihypertensive treatment and 24-hour MAP. IVS: interventricular septum; LAVI: left atrial volume index; LVIDd: left ventricular internal diameter in diastole; LVIDs: left ventricular internal diameter in systole; LVMI: left ventricular mass index; LVPW: left ventricle posterior wall; MAP: mean arterial pressure; PASP: pulmonary artery systolic pressure; RVIDd: right ventricular internal diameter in diastole; RWT: relative wall thickness; TAPSE: tricuspid annular plane systolic excursion.*

## Discussion

The findings of this cohort study suggest that non-dipping population had a higher prevalence of cardiovascular events, especially hypertension, heart failure and coronary heart disease. Previous reports have shown this relationship. The OPERA (Oulu Project Elucidating Risk of Atherosclerosis) population-based cohort study evaluated risk factors and endpoints of atherosclerotic cardiovascular disease during a 21-year follow-up. It showed that a non-dipping pattern was associated with non-fatal cardiovascular events in the long term and that this effect was independent of the conventional risk factors including office and ambulatory BP levels.^21^

Regardless of its exact contribution, the non-dipping blood pressure profile both in the overall and hypertensive patient population demonstrates statistical correlation with a higher AASI. In previous research, a reduced blood pressure dipping pattern was linked to higher AASI and measures of vascular inflammation. Systolic blood pressure dipping percentage, AASI, monocyte-lymphocyte ratio (MLR) and neutrophils-lymphocyte ratio (NLR) were associated with major adverse cardiovascular events (MACE) on univariate analyses. Increased NLRs were independently associated with MACE and MLR with cardiovascular death or non-fatal stroke. This information endorses the association between AASI, inflammation and cardiovascular risk.^19^

Regarding the echocardiographic variables, a higher LVMI, especially in men, LVPW thickness and LVIDd was found in the non-dipping group, resulting in a higher frequency of eccentric hypertrophy. Functionally, there seems to be no clinically significant difference of systolic function of the left ventricle. However, in the non-dipping group, a higher frequency of diastolic dysfunction suggests a higher likelihood of alterations in ventricular relaxation.

This is in agreement with a meta-analysis of 3,591 untreated adults (1,291 non-dipping and 2,300 dipping hypertensives), where LVMI, RWT, and LAD were greater in non-dipping hypertensives than their dipping counterparts, while the E/A ratio was reduced. Consistent with the findings of this meta-analysis, our study revealed a notable association of a non-dipping profile with diastolic dysfunction, particularly Grade I. This link underscores the importance of non-dipping pattern’s role in predicting adverse cardiac remodelling as a critical manifestation of target organ damage.^22^

While this study establishes a distinct association between a non-dipping profile and adverse cardiac remodelling in both hypertensive and non-hypertensive individuals, additional research is required to assess the impact of this pattern on future cardiovascular events. Moreover, it is necessary to explore whether reversing the non-dipping pattern could mitigate adverse cardiac remodelling. Such findings could have significant implications for therapeutic strategies and patient outcomes.

## Limitations

This retrospective cohort study, investigating the association of a non-dipping profile with echocardiographic changes, encounters several potential limitations. Firstly, the temporal disconnection between the 24-hour ambulatory blood pressure monitoring (ABPM) and the echocardiograms, which were conducted up to two years apart, may introduce temporal bias, limiting our ability to establish a causal relationship between non-dipping status and cardiac alterations. Secondly, reliance on medical records for data collection could lead to information bias due to variability in the quality and completeness of the records. Additionally, since ABPM and echocardiography were not performed simultaneously for all subjects, there could be intervening variables and lifestyle changes affecting both blood pressure and cardiac structure and function that are not accounted for in the study. Finally, the study’s retrospective design inherently limits the control over confounding variables and potential selection bias, which might influence the generalizability of the findings. Addressing these issues in future prospective studies could help clarify the impact of a non-dipping BP profile on cardiac structure and function.

## Conclusions

A non-dipping profile is clearly linked with both structural and functional echocardiographic alterations, as well as adverse cardiac remodelling in hypertensive and non-hypertensive individuals. However, the feasibility and potential benefits of reversing a non-dipping profile to mitigate these adverse effects remain uncertain. Further investigation is necessary to determine if such an intervention could be a viable therapeutic approach to improve cardiac outcomes.

## Perspectives

Our study highlights the significant impact of non-dipping blood pressure patterns on cardiac structure and function, underscoring the potential of these patterns as independent predictors of adverse cardiac remodeling. The findings suggest that individuals with non-dipping BP profiles, regardless of hypertension status, exhibit an increased left ventricular mass index, left ventricular internal diameter in diastole, and prevalence of diastolic dysfunction. These results have broad implications for the management of cardiovascular risk in both hypertensive and normotensive populations. Future research should focus on longitudinal studies to establish causality and explore interventions aimed at modifying BP dipping patterns.

Additionally, expanding this research to diverse populations can enhance the generalizability of these findings. The potential to use non-dipping BP profiles as a marker for early intervention in cardiovascular care could impact current practices and improve patient outcomes.

## Novelty and Relevance

### What Is New?

- Identification of non-dipping BP patterns as independent predictors of adverse cardiac remodeling.
- Detailed echocardiographic characterization of cardiac changes associated with non-dipping BP profiles.
- Inclusion of both hypertensive and normotensive individuals in a large cohort study.

### What Is Relevant?

- Demonstrates the importance of monitoring BP patterns beyond traditional daytime measurements.
- Highlights the association between non-dipping BP profiles and increased structural and functional cardiac changes.
- Provides evidence to support the use of 24-hour ambulatory blood pressure monitoring in routine clinical practice.

### Clinical/Pathophysiological Implications

- Non-dipping BP profiles can serve as early indicators of adverse cardiac remodeling, prompting timely intervention.
- Potential to refine risk stratification in patients with hypertension and those at risk for cardiovascular diseases.
- Emphasizes the need for targeted therapeutic strategies to address abnormal BP patterns, which could mitigate the progression of cardiac dysfunction and improve long-term outcomes.

## Data Availability

All data referred to in this manuscript are available at a public repository online (Synapse). The data supporting the findings of this study can be accessed at https://doi.org/10.7303/syn59775014. For further inquiries, please contact the corresponding authors, Dr. Federico Ramos at and Dr. Daniel Villarreal at. Data access is subject to compliance with local and international regulations governing the use of medical records and participant confidentiality.

## Acknowledgements

We express our gratitude to Dr. Cindy Restrepo, Cardiology Fellow at Universidad del Bosque, Dr. Andrés Felipe Barrera, general physician and Juan David Castillo, a medical student at Universidad de los Andes, for their invaluable assistance during the data collection phase of this study. We also extend our appreciation to the Department of Internal Medicine and the Department of Cardiology at Hospital Universitario Fundación Santa Fe de Bogotá, as well as the Faculty of Medicine at Universidad de los Andes, Bogotá, Colombia, for their unwavering support throughout the development of this study.

## Non-standard Abbreviations and Acronyms

AASI: Ambulatory Arterial Stiffness Index
ABPM: Ambulatory Blood Pressure Monitoring
ANCOVA: Analysis of Covariance
ASE: American Society of Echocardiography
BMI: Body Mass Index
BP: Blood Pressure.
IRB: Institutional Review Board
IRT: Isovolumetric Relaxation Time
LAD: Left Atrial Diameter
LVEF: Left Ventricular Ejection Fraction
LVPW: Left Ventricular Posterior Wall
LVMI: Left Ventricular Mass Index
LVIDd: Left Ventricular Internal Diameter in Diastole
LVIDs: Left Ventricular Internal Diameter in Systole
MACE: Major Adverse Cardiovascular Events
MPI: Myocardial Performance Index
MLR: Monocyte-Lymphocyte Ratio
NLR: Neutrophil-Lymphocyte Ratio
OPERA: Oulu Project Elucidating Risk of Atherosclerosis
RWT: Relative Wall Thickness

## Sources of Founding

This research project was funded by Fundación Santa Fe de Bogotá.

## Supplemental Material

Supplemental Methods

Supplemental Results Tables S1 – S2

Major Resource Table

